# The Seattle Flu Study: a multi-arm community-based prospective study protocol for assessing influenza prevalence, transmission, and genomic epidemiology

**DOI:** 10.1101/2020.03.02.20029595

**Authors:** Seattle Flu Study Investigators, Helen Y. Chu, Michael Boeckh, Janet A. Englund, Michael Famulare, Barry R. Lutz, Deborah A. Nickerson, Mark J. Rieder, Lea M. Starita, Jay Shendure, Trevor Bedford, Co-Investigators, Amanda Adler, Elisabeth Brandstetter, Chris D. Frazar, Peter D. Han, Reena K. Gulati, James Hadfield, Michael L. Jackson, Anahita Kiavand, Louise E. Kimball, Kirsten Lacombe, Jennifer Logue, Victoria Lyon, Kira L. Newman, Thomas R. Sibley, Monica L. Zigman Suchsland, Caitlin Wolf

## Abstract

**Introduction:** Influenza epidemics and pandemics cause significant morbidity and mortality. An effective response to a potential pandemic requires the infrastructure to rapidly detect, characterize, and potentially contain new and emerging influenza strains at a population level. The objective of this study is to use data gathered simultaneously from community and hospital sites to develop a model of how influenza enters and spreads in a population.

**Methods and Analysis:** Starting in the 2018-19 season, we have been enrolling individuals with acute respiratory illness from community sites throughout the Seattle metropolitan area, including clinics, childcare facilities, Seattle-Tacoma International Airport, workplaces, college campuses, and homeless shelters. At these sites, we collect clinical data and mid-nasal swabs from individuals with at least two acute respiratory symptoms. Additionally, we collect residual nasal swabs and data from individuals who seek care for respiratory symptoms at four regional hospitals. Samples are tested using a multiplex molecular assay, and influenza whole genome sequencing is performed for samples with influenza detected. Geospatial mapping and computational modeling platforms are in development to characterize the regional spread of influenza and other respiratory pathogens.

**Ethics and Dissemination:** The study was approved by the University of Washington’s Institutional Review Board. Results will be disseminated through talks at conferences, peer-reviewed publications, and on the study website (www.seattleflu.org).

**Article Summary:** *Strengths and limitations of this study:*

- Large-scale multiple-arm study of respiratory illness characterization with collection of samples from individuals in the community as well as in ambulatory care and hospital settings
- Integration of sociodemographic, clinical, and geospatial data on a regional level
- Multiplex molecular testing for multiple viral and bacterial pathogens and whole genome sequencing of influenza for detailed molecular epidemiologic characterization and transmission mapping
- Geographically and socioeconomically diverse sampling of community-based acute respiratory illnesses

## Introduction

In the United States, annual influenza epidemics cause 9-45 million illnesses, 140,000-810,000 hospitalizations, and 12,000-67,000 deaths. The 1918 pandemic, the worst for which reliable records exist, resulted in an estimated 675,000 deaths in the United States and 50 million worldwide (1-3). More recently, the 2009 H1N1 pandemic caused 12,500 deaths and 60.8 million infections in the United States, with an estimated global burden of 284,500 deaths (4, 5).

Active surveillance for influenza is essential to monitor the impact of seasonal influenza and to detect and characterize emerging influenza viruses. In the United States, surveillance systems rely primarily on medically-attended illnesses. This underestimates true influenza-related disease burden by 50% or more (6). Individuals may not seek care, diagnostic tests are often underutilized in the community, and hospitalizations may be attributed to chronic conditions exacerbated by influenza (7, 8). Furthermore, recent studies have estimated that 20% of individuals with influenza infection are asymptomatic but contribute to community transmission (9-11).

Community-wide studies provide a mechanism to identify influenza among individuals who do not present for care and who may provide the first signal of an impending pandemic (12). Previous community-wide studies of influenza, including family-based prospective studies, conducted in the 20^th^ century, have provided important data on transmission dynamics, including the role of households and schools in driving seasonal epidemics. More recent prospective studies employing molecular detection methods have highlighted the role of asymptomatic individuals in disease transmission (13). However, a common limitation of community-based studies is they do not routinely integrate their data with inpatient hospital and ambulatory care surveillance to understand the transmission and burden of influenza at a population level within a specific geographic area.

Despite substantial progress in the estimation of the burden of influenza through large-scale surveillance studies, there is an ongoing need for improved near-real-time monitoring that is coupled to pandemic preparedness. We envision that pandemic control will require the rapid accrual of actionable information from a diversity of sources. For example, rapid genome sequencing of influenza strains throughout a city could feed the creation of actionable maps that identify new and emerging influenza strains their transmission dynamics. With such a system in place, we can more effectively develop and test strategies to rapidly deploy vaccines, antivirals, and non-pharmaceutical interventions to areas with early influenza detection. This combination of rapid detection and deployment of interventions may provide a new paradigm to contain outbreaks.

This manuscript describes the protocol for the Seattle Flu Study (SFS), a multi-armed regional study of influenza at a city-wide scale that integrates community, ambulatory care, and inpatient surveillance at unprecedented intensity. The objectives of this study are to gather data from community and hospital sites to advance our understanding of how influenza enters and spreads in a population, as well as to create a city-wide platform for testing novel interventions that may limit or contain influenza outbreaks.

### Aims and Hypothesis

The primary aim of this study is to conduct a community-wide prospective study of symptomatic influenza prevalence, transmission, and genetics. Our approach combines cross-sectional surveillance methodologies across diverse prospective cohorts with streamlined data integration to drive near-real-time analysis to guide disease control activities.

The primary hypothesis is that a large-scale cross-sectional design can detect early cases of influenza, monitor community-level transmission through viral genetic and spatial-demographic analysis, and be used to inform public health and clinical interventions.

## Methods and Analysis

### Study design

The Seattle Flu Study is a multi-armed surveillance study. The arms include (1) community cross-sectional, (2) clinical cross-sectional, (3) prospective clinical cohort, (4) prospective childcare cohort, and (5) active clinical surveillance. This protocol describes the Seattle Flu Study as implemented in Year 1. Sub-studies and modifications in future years will be described in subsequent publications.

### Study overview and setting

In this study, participants are eligible if they have two or more acute respiratory illness (ARI)-associated symptoms (**Table 1**) or a medically-collected respiratory specimen. The presence of a subjective or objective fever is not required. All participants have a respiratory specimen collected that is linked with demographic information, illness characteristics, behavioral, and other clinical metadata. The primary outcome is influenza infection status as defined by molecular diagnostic testing. The study takes place during influenza season (October to May) annually, starting with 2018-2019.

**Table 1.** Acute respiratory illness symptoms used to trigger the collection of a mid-nasal swab in the community cross-sectional, prospective clinical cohort, or childcare cohort arms of the Seattle Flu Study. Study participants must have at least two self-reported symptoms in the 7 days prior to swab collection.

|  |  |  |  |
| --- | --- | --- | --- |
| Fever | Cough | Sore throat | Headache |
| Diarrhea | Nausea or vomiting | Runny or stuffy nose | Rash |
| Fatigue (tiredness) | Muscle or body aches | Increased trouble with breathing | Ear pain or discharge |

The study occurs in a variety of settings throughout the Seattle metropolitan area (Figure 3). Participants in the community cross-sectional arm are recruited at standalone kiosks in public settings including clinical facilities, university campuses, airports, workplaces, homeless shelters, and high-traffic tourist areas. Participants in the clinical cross-sectional and prospective clinical cohort arms are recruited from traditional sites of care such as hospitals and medical clinics. Participants in the prospective childcare cohort arm are recruited from childcare facilities. All collection sites are located in the greater Seattle metropolitan area, including King, Pierce, Snohomish, Skagit, and Island Counties, Washington, USA.

The community cross-sectional arm enrolls individuals with two or more protocol-defined respiratory symptoms (Table 1). One mid-nasal swab is collected at the time of enrollment and linked with questionnaire data. These individuals do not have follow up. Individuals are eligible to re-enroll for new ARI episodes every 14 days. Surface and bioaerosol samples are additionally collected at a subset of sites and tested for respiratory viruses using the same laboratory methods as the respiratory specimens.

The clinical cross-sectional arm uses residual clinically-collected respiratory specimens of all types from participating hospitals and clinics, regardless of clinical test result. Specimens are aliquoted and undergo the same laboratory processing as above. Demographic and clinical metadata are extracted from the electronic medical record (EMR) extraction and linked to the corresponding respiratory specimen. There is no direct contact with these individuals.

The prospective clinical cohort enrolls hospitalized individuals with a lab-confirmed respiratory virus as tested by the hospital’s clinical lab. Participants are enrolled by the same method as the community cross-sectional arm. They are surveilled and sampled daily for up to seven days after diagnosis.

The prospective childcare cohort enrolls children attending participating daycares. Children are enrolled prior to the local influenza season, and a baseline mid-nasal swab is collected at this time. They are then surveyed weekly for the duration of the local influenza season for development of ARI symptoms. If symptom criteria are met, an additional mid-nasal swab and corresponding data are collected.

The active clinical surveillance arm utilizes specimens collected at participating medical clinics as well as through the Washington site of the United States Influenza Vaccine Effectiveness (US Flu VE Network), details of which have been previously published (14, 15). In the US Flu VE Network, patients seeking ambulatory care for ARI are prospectively identified and recruited into the study, with collection of clinical data and a nasal swab. Clinical data and residual nasal swabs from individuals enrolled in the Flu VE study were further analyzed for the Seattle Flu Study.

### Study population inclusion/exclusion criteria

Participants are eligible for inclusion if they meet all the following criteria for their study arm and do not meet any exclusion criteria. Prospectively recruited participants are excluded from the study if they are unable to provide consent themselves or through a legally authorized representative (LAR) or if they are incarcerated, wards of the state, or have any condition that, at the investigators’ discretion, may preclude or limit participation with study procedures.

Participants in the community cross-sectional study arm are eligible at any age, and can enroll at any of the kiosks if they have two or more new or worsening ARI symptoms and are able to provide consent themselves or through an LAR. In addition to overall study criteria, individuals are excluded if they have previously enrolled into the study within 14 days. For individuals under age 18, consent is obtained from an LAR.

There is no participant interaction in the clinical cross-sectional study arm. Data is utilized if they have a respiratory specimen collected at clinician discretion from a participating site within the five-county study surveillance region during the study period.

Prospective clinical cohort participants are eligible if they are at least 18 years old, inpatient at a participating hospital, English- or Spanish-speaking, have a laboratory-confirmed respiratory virus infection, and can consent and sign HIPAA authorization for themselves or through an LAR.

Prospective childcare cohort participants are eligible if they are children attending a participating childcare facility and have an LAR to provide consent.

Patients are eligible for US Flu VE Network enrollment if they are age 6 months or older as of September 1st 2018, have a cough of <8 days’ duration, and have not used an antiviral medication in the past 7 days.

### Consent and recruitment

This study is approved by the University of Washington Human Subjects Institutional Review Board. Recruitment methods vary by study arm. The community cross-sectional, prospective clinical cohort, and prospective childcare cohort arms recruit participants by actively approaching potentially eligible individuals, and through digital media and flyers. For all three study arms, eligible and interested participants are consented using an electronic consent form. For consent details by participant age, see **Figure 1**. In accordance with IRB approval, consent and Health Insurance Portability and Accountability Act (HIPAA) authorizations for participants in the clinical cross-sectional study are waived.

**Figure 1.**
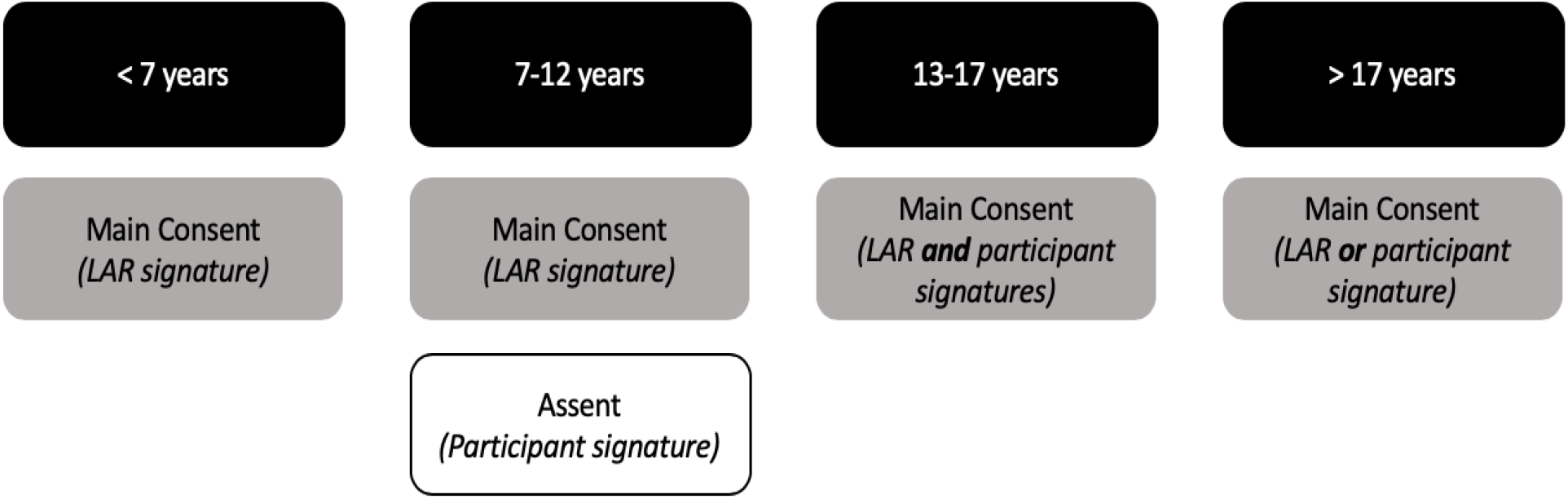
Documentation of written, informed consent by age of participant in the community cross-sectional, and prospective clinical and childcare cohorts of the Seattle Flu Study. If the participant is unable to provide informed consent due to cognitive impairment or because they have not attained the legal age for consent, a legally-authorized representative (LAR) may sign the consent form on their behalf. Participants enrolled by Seattle Children’s Hospital staff or participants enrolled into the prospective clinical cohort sign a HIPAA Agreement in addition to the main consent.

### Sample size

The goal of the study is to obtain samples and data from cases of influenza in the Seattle metropolitan area. In 2017-2018, there were 6953 positive tests for influenza in Washington state that were reported to the Centers for Disease Control through participating clinical laboratories, which only represent a subset of the total cases diagnosed (16, 17).

### Outcomes

#### Primary outcome

- Clinical and sociodemographic characteristics of individuals with ARI attributable to influenza
- Geospatial mapping of influenza ARI cases
- Evaluation of genetic diversity of circulating influenza strains in the Seattle metropolitan area, by variables including age, vaccine status, site of collection, and home census tract

#### Secondary outcomes

- Prevalence of ARI specifically attributable to influenza, as well as to RSV, adenovirus, coronavirus, rhinovirus and other respiratory pathogens
- Clinical, geospatial, and sociodemographic characteristics of participants with ARI respiratory pathogens other than influenza
- Geospatial mapping of ARI cases attributable to other respiratory pathogens
- Genetic diversity of circulating strains of respiratory pathogens in the Seattle metropolitan area, by age, vaccine status, site of collection, and home census tract
- Impact of bacterial co-detection on viral ARI disease severity and outcomes
- Molecular epidemiology and transmission dynamics of respiratory pathogens within sites of collection
- Impact of viral co-infection on viral ARI disease severity and outcomes
- Prevalence and concordance of environmental sampling by sample type and sampling location
- Viral kinetics of respiratory pathogens among individuals with longitudinal sample collection
- Prevalence and predictors of care-seeking and influenza vaccine receipt within participants
- Viral load and its relationship with clinical disease characteristics in both single and multiple virus infections
- Correlation of viral load with molecular markers of nasal swab sampling efficiency
- Probability of influenza based on individual-level factors, including symptoms

### Data collection methods

For participants enrolled in the clinical cross-sectional or prospective clinical arms, EMR data are obtained through the clinical data warehouse, which consolidates patient-level medical data from multiple sources. Survey data from eligible and enrolled participants in the community cross-sectional and prospective arms are collected through the FluTrack app (Audere, Seattle, WA), a mobile-enabled app created for the Seattle Flu Study and administered on a tablet. For participants unable or unfamiliar with the use of the tablet, questionnaires on the FluTrack app are administered by study staff verbally in English or Spanish. If the participant is under 7 years old, their LAR completes the questionnaire on their behalf. For participants aged 7-12 years, the LAR may decide whether to complete the survey on the participant’s behalf or in collaboration. Participants aged 13 years or older complete the questionnaire. Data are collected on US Flu VE Network enrollees via interview and extraction from EMR and administrative healthcare databases.

### Participant retention and compensation

Prospective clinical cohort participants receive a gift card for each longitudinal study specimen that is collected (nasal swabs and blood). Participants in the community cross-sectional arm receive a gift card for completing the study. Participants in the prospective childcare cohort receive a gift card for each episode with collection of a mid-nasal swab. There is no remuneration for participants in the clinical cross-sectional arm since there is no participant interaction.

### Data security and storage

All information from the study subjects is kept confidential. All forms and specimens are assigned a participant identification number, given to the participant upon enrollment in the study, that will be used in the place of names whenever possible. Data will be collected electronically in a Title 21 CFR Part 11 compliant, password protected and auditable database. The list linking the participant to the ID number will be stored separately from the database. Access to identifiable information are limited to the study staff. Electronic files are secured via logon password protection for study accounts. Any datasets that include identifiable information are stored in a HIPAA-compliant manner. No identifying information is included on any data sent to the broader study team or any other data-sharing repositories. All data files transferred for this study will be transferred via encrypted software and the original files are kept on the server at the Fred Hutchinson Cancer Research Institute in a database.

Identifiable numbers are kept on biospecimens (nasal swabs, environmental samples) while the laboratory processing occurs. All subjects who consent to participate are asked to approve the storage of their biospecimens (nasal swabs). Backup aliquots are kept until all laboratory assays are completed with adequate quality control. Persons who consent to the trial, but who do not want their biospecimens stored may still participate in the trial. Their biospecimens are tested as per protocol, but the remaining aliquots will be destroyed.

Identifiers are kept on all data files until the study is closed out. Primary data collection sources will be kept for at least 5 years following the publication of the primary result from this study.

Once this time elapses and the electronic data files are fully cleaned, any paper forms will be destroyed.

### Data quality

Data are checked for missing or unusual values and checked for consistency in the centralized data capture system. Computerized checks are conducted weekly to identify missing, inconsistent or out of range data. Any suspect data is raised as data queries, and are investigated by the study coordinators.

### Protection against risks

All identifying data is stored using standard security techniques. Hard copies of data collection materials that have identifiers will be locked in the office of the study PI or a room with limited access by specific individuals. When possible, redacted (de-identified) versions of the data will be used for coding and data analysis. Personal identifiers will be stored in the database on a password protected network that is HIPAA-compliant, and only accessible to specific individuals. Transfer or storage on portable devices (e.g., laptops, flashdrives) will be encrypted. The devices on which this information is stored is accessible only to individuals who need access to the data.

### Specimen collection methods

#### Respiratory specimens

Prospectively collected respiratory specimens are collected using a sterile Copan flocked respiratory swab inserted and rotated mid-nasally. Respiratory specimens are transported to the research laboratory in Universal Viral Transport Medium (UTM, Becton, Dickinson and Company, Franklin, NJ). Clinically-collected respiratory specimens are obtained from each participating hospital or clinic laboratory, maintained at 4C, and transported to the research laboratory within 3-7 days of specimen collection on wet ice (Figure 2).

**Figure 2.**
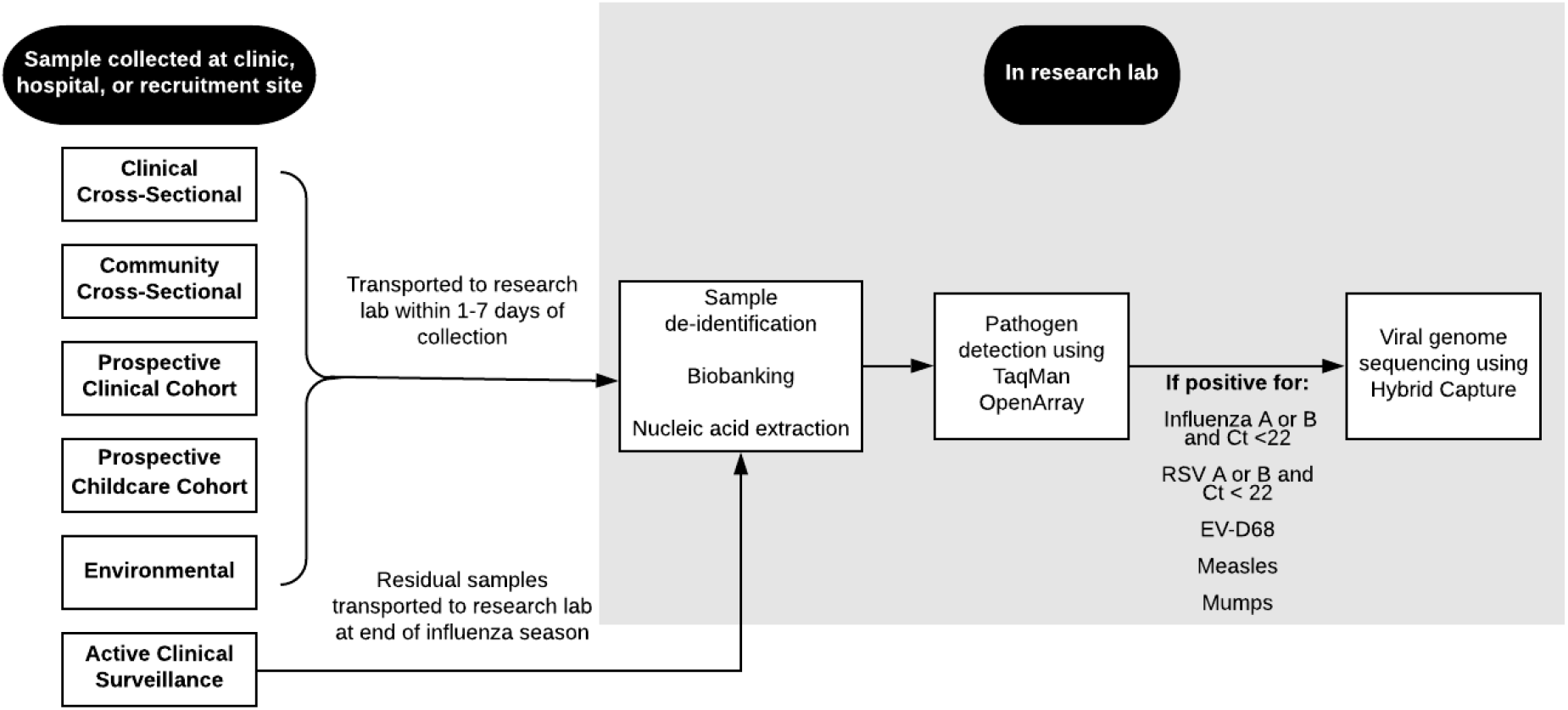
Laboratory pipeline for samples included in the Seattle Flu Study from time of collection through sequencing. Abbreviations: Ct, cycle threshold; EV-D68, enterovirus D68.

**Figure 3.**
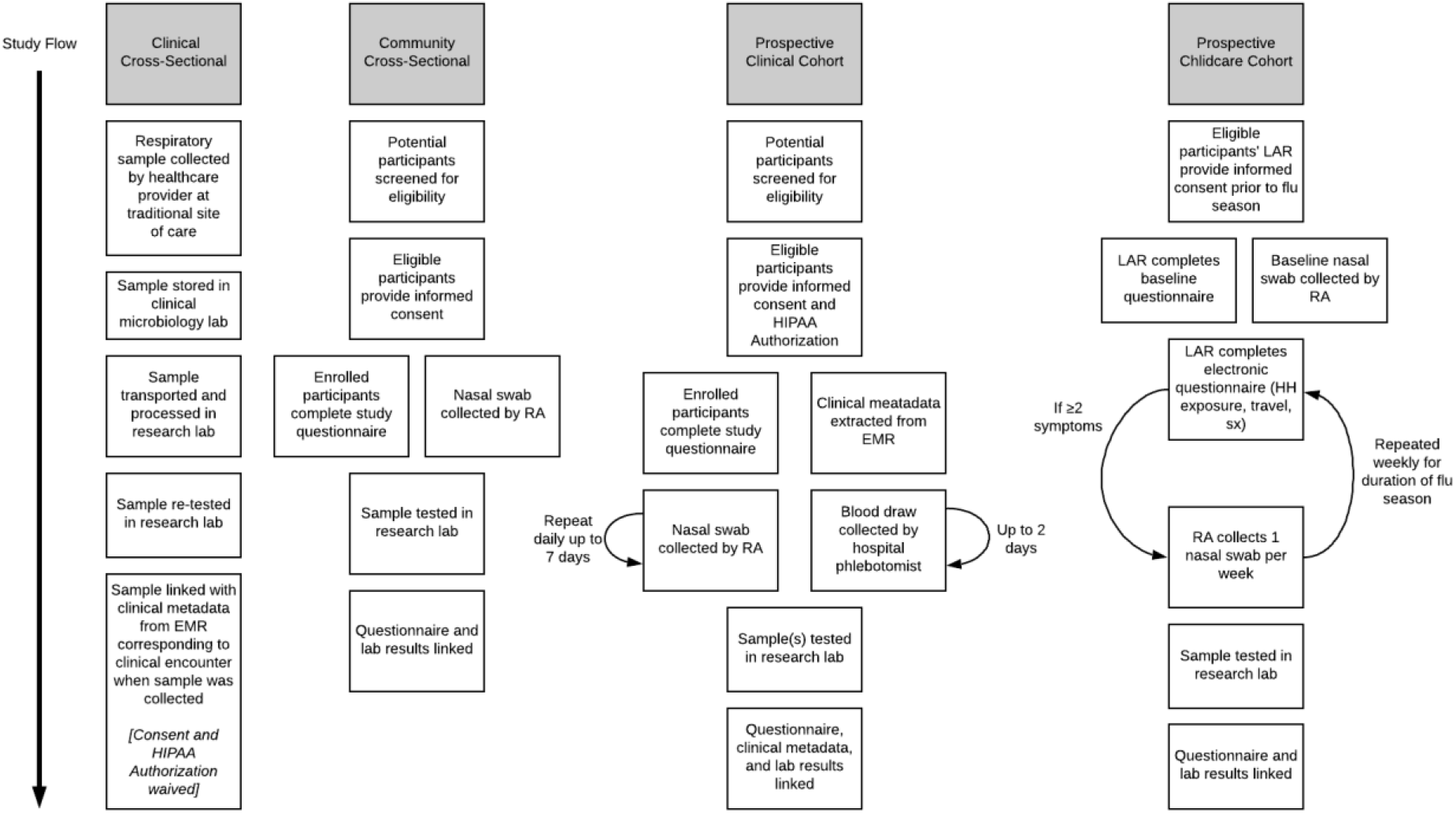
Outline of participant flow for different arms of the Seattle Flu Study. Abbreviations: EMR, electronic medical record; HH, household; RA, research assistant; LAR, legally authorized representative; sx, symptoms. (Note: active clinical surveillance arm not included in figure. For methods, see [13, 14].)

#### Environmental samples

Both bioaerosol and surface samples are collected longitudinally at select community cross-sectional and prospective childcare cohort sites. Prespecified high-touch surfaces are sampled by wiping a designated object or 10 cm^2^ area with a synthetic polyester swab and transported to the lab in UTM. Bioaerosol samples are collected over a 90-minute sampling period using an SKC QuickTake 30 Air Pump at prespecified locations with high foot traffic and low air ventilation at select community sites. These samples are collected on membrane filter paper inside the air pump. Environmental samples undergo the specimen handling procedures as above.

### Laboratory methods

All study respiratory specimens are aliquoted in triplicate and barcoded using a unique identifier that can be linked back to the participant or site of collection. Samples were frozen at −80°C until thawed for extraction (Figure 2). Total nucleic acids are extracted from 200 µl of UTM using Magna Pure 96 small total nucleic acids extraction kit (Roche). Extracted nucleic acids are screened for the presence of respiratory pathogens by TaqMan RT-PCR on the OpenArray platform (Thermo) (Table 2). Specimens that test positive for influenza by RT-PCR are then sequenced using a modified oligo capture protocol. Total RNA is converted to cDNA and sequencing libraries are constructed using the Illumina TruSeq RNA Library Prep for Enrichment kit, as outlined by the manufacturer (Illumina, San Diego, CA). Library samples are ranked by TaqMan CRT value, and 24 samples with similar CRT values are pooled prior to oligo capture. CRT values are further used to determine the viral load of each sample and inform the fraction of each capture pool that a given sample represents. Each pool of flu-positive samples is hybridized overnight to a custom pool of capture oligonucleotides following the manufacturer’s recommended protocol (Twist Bioscience, San Francisco, CA). Final libraries are sequenced on the Illumina Miseq, NextSeq or NovaSeq platform with paired end 150bp reads.

**Table 2.** Pathogens for which all Seattle Flu Study respiratory specimens are tested using a TaqMan RT-PCR.

| Viruses | Bacteria |
| --- | --- |
| Influenza A—H3N2 | *Streptococcus pneumoniae* |
| Influenza A—H1N1 | *Mycoplasma pneumoniae* |
| Influenza A—Pan | *Chlamydia pneumoniae* |
| Influenza B | *Bordetella pertussis*<sup>1</sup> |
| Influenza C |  |
| Respiratory syncytial viruses A and B |  |
| Parainfluenza viruses 1-4 |  |
| Coronavirus 229E, NL63, OC43, and HKU1 |  |
| Adenovirus |  |
| Rhinovirus |  |
| Measles |  |
| Mumps |  |
| Human metapneumovirus |  |
| Human parechovirus |  |
| Enterovirus<sup>2</sup> |  |
| Enterovirus D68 |  |
| Human Bocavirus |  |

<sup>1</sup> Includes *Bordetella bronchiseptica* and *Bordetella parapertussis*
<sup>2</sup> All enterovirus species A, B, C, D, and G, including: all Coxsackie serotypes under species A, B, C; all Echovirus serotypes; all Poliovirus serotypes (1-3).

### Statistical methods

Prevalence of respiratory pathogens are analyzed as the number of cases detected out of the total number of episodes with testing. For influenza-specific analyses, prevalence is defined as the number of cases of influenza out of the total number of episodes with testing. Risk ratios and associated 95% confidence intervals are estimated for the cross-sectional study arms using Poisson or negative binomial regression. Similarly, risk ratios and associated 95% confidence intervals are estimated for the prospective clinical and childcare cohorts. For questionnaire data, descriptive statistics are calculated and association with respiratory pathogen prevalence analyzed using parametric and nonparametric tests, as appropriate given the distributions. Geospatial incidence maps are estimated using generalized additive mixed models with spatial, temporal, age, and non-structured random effects (18). Phylogenetic analyses are based on the Nextstrain pipeline (19).

For longitudinal data such as viral kinetics, we are conducting a descriptive analysis of group means over time and using mixed models, general estimating equations, and quantile regression to assess for change over time, adjusting for potential confounders, including age, comorbidities, and vaccination status.

### Missing data

For questionnaire data in the cross-sectional study, we do not anticipate substantial missing data because completion of the survey is a criterion for participation, though participants can state that they prefer not to answer specific questions. For longitudinal data, we are using complete case analysis. For analyses and subgroups where complete case analysis leads to loss of 10% or more of subjects, we are performing multiple imputation and sensitivity analyses.

### Subgroup analyses

The main subgroups of interest are age, recruitment site (homeless shelter, childcare, clinical, other) and vaccination status.

## Ethics and Dissemination

### Monitoring

The Seattle Flu Study has a scientific advisory board to which it reports bi-annually. These reports include updates on enrollment, preliminary results, sub-studies, and protocol modifications.

### Assessment of harms and adverse events

Participants receive contact information from the study team and are encouraged to contact the team if they experience any participation-related. Given the minimally-invasive nature of the sample collection procedure, harms and adverse events are very unlikely. Should any significant adverse events arise related to the study procedures, they will be reported to institutional authorities at University of Washington and any applicable participating site.

### Informed consent

All participants in the community cross-sectional and prospective arms are informed about the study purpose, risks, and benefits, and they or their LAR provide written informed consent via an electronic consent form prior to study participation. US Flu VE Network enrollees provide informed verbal consent for respiratory swab collection and informed written consent if providing blood. Participants are informed during the consent process that they may withdraw from the study at any time.

### Access to data

The data will be accessed only by authorized individuals on the study team. Access to de-identified, aggregated data and analysis code will be publicly available on the study web page (www.seattleflu.org).

### Dissemination plans

Our study group will present the results at national and international research conferences and through peer-reviewed publications and on the study website (www.SeattleFlu.org). Any changes to the study protocol will be communicated to journals in accordance with that journal’s reporting policies. We will follow STrengthening the Reporting of OBservational studies in

Epidemiology (STROBE) reporting guidelines, as applicable (20)]. Study materials, including site-specific questionnaires and FluTracker app screenshots, will be available upon request with approval of the study team.

### Patient and public involvement

Most public involvement is as research participants. There are no current plans to involve participants in the study recruitment, conduct, analysis, or dissemination, though aggregate data will be made publicly available.

## Discussion

We present the study design for large-scale assessment of the burden of ARI attributable to influenza and other respiratory pathogens to map the clinical and molecular epidemiology of influenza in a major metropolitan area. This will provide the framework for further studies of strategies for rapid, accurate identification and deployment of interventions to interrupt spread of infectious disease in major metropolitan areas.

## Data Availability

The data will be accessed only by authorized individuals on the study team. Access to deidentified, aggregated data and analysis code will be publicly available on the study web page (www.seattleflu.org).

## Funding Statement

The Seattle Flu Study is funded through the Brotman Baty Institute. The funder was not involved in the design of the study, does not have any ownership over the management and conduct of the study, the data, or the rights to publish.

## Authors Contributions

Helen Y. Chu, Michael Boeckh, Janet A. Englund, Michael Famulare, Barry R. Lutz, Deborah A. Nickerson, Mark J. Rieder, Lea M. Starita, Jay Shendure, and Trevor Bedford, designed and implemented the protocol, they also wrote, reviewed and edited the written protocol. Elisabeth Brandstetter, Jennifer Logue, Kira L Newman, and Caitlin Wolf contributed to the design of the prospective clinical and cross-sectional community study arms and also wrote, reviewed, and edited the written protocol. Reena K. Gulati, Louise E. Kimball, Michael L. Jackson assisted in the design of the overall protocol. Amanda Adler and Kirsten Lacombe contributed to the design of the prospective childcare cohort and reviewed and edited the written protocol. Thomas R. Sibley, James Hadfield, Peter D. Han, Chris D. Frazar, and Anahita Kiavand contributed to the development of the laboratory and data processing aspects of the protocol and reviewed and edited the written protocol, Monica L. Zigman Suchsland and Victoria Lyon contributed to the clinical data aspect of the protocol and reviewed and edited the written protocol.

## Competing Interests

Amanda Adler, Elisabeth Brandstetter, Michael Famulare, Chris D. Frazar, Peter D. Han, Reena K. Gulati, James Hadfield, Michael L. Jackson, Anahita Kiavand, Louise E. Kimball, Kirsten Lacombe, Jennifer Logue, Victoria Lyon, Kira L. Newman, Thomas R. Sibley, Jay Shendure, Lea Starita, Monica L. Zigman Suchsland, and Caitlin Wolf declare no competing interests. Helen Y Chu receives research support from Sanofi, Cepheid, and Genentech/Roche and is a consultant for Merck. Janet Englund receives research support to her institution from Astrazeneca, GlaxoSmithKline, Merck, and Novavax and is a consultant for Sanofi Pasteur and Meissa Vaccines.

